# Diagnostic technology for COVID-19: comparative evaluation of antigen and serology-based SARS-CoV-2 immunoassays, and contact tracing solutions for potential use as at-home products

**DOI:** 10.1101/2020.06.25.20140236

**Authors:** Mehdi Jorfi, Nell Meosky Luo, Aditi Hazra, Fanny Herisson, Glenn Miller, James A. Toombs, David R. Walt, Paolo Bonato, Rushdy Ahmad, COVID-19 Direct-to-Consumer Task Force Investigators

## Abstract

As the United States prepares to return to work and open up the economy in the midst of the COVID-19 pandemic without an available vaccine or effective therapy, testing and contact tracing are essential to contain and limit the spread of the COVID-19 virus. In response to the urgent public health need for accurate, effective, low-cost, and scalable COVID-19 testing technology, we evaluated and identified diagnostic solutions with potential for use as an at-home product. We conducted a deep horizon scan for antigen and serology-based diagnostics and down-selected to the most promising technologies. A total of 303 candidate products (138 antibody and 44 antigen tests) were identified. Product evaluations were based entirely on company-provided data. 73 serology-based antibody tests passing an initial scoring algorithm based on specificity and sensitivity data were then further evaluated using a second scoring algorithm. This second algorithm included a review of additional technical specifications of the devices, an analysis of supply chain, manufacturing, and distribution capacity of each vendor. 24 potential antibody products met the selection criteria for further direct laboratory evaluation. The performance metrics for selection of these 24 products are currently being evaluated in a Mass General Brigham laboratory. Testing alone might not be sufficient to prevent the spread of a highly contagious disease like COVID-19. Manual contact tracing could complement testing, but it is likely to fail in identifying many individuals who were in contact with a given COVID patient. The proliferation of smartphones in the population has enabled the development of solutions that can provide public health officials with valuable information for rapid and accurate contact tracing. Besides, electronic-based contact tracing solutions can be augmented by symptom self-reports gathered using electronic patient reported outcome (ePRO) platforms and by physiological data collected using wearable sensors. We performed a detailed assessment of 12 ePRO solutions, 27 wearable sensors, and 44 electronic-based contact tracing solutions. These technologies were evaluated using criteria developed to assess their suitability to address the COVID-19 pandemic. We identified a number of solutions that could augment if not provide a more effective alternative to manual contact tracing. Finally, we propose a theoretical framework in which ePRO platforms, wearable sensors, and electronic-based contact tracing solutions would be utilized in combination with molecular and serological tests to identify and isolate COVID-19 cases rapidly.

## Introduction

The spread of the novel severe acute respiratory syndrome coronavirus, SARS-CoV-2, which originated from Wuhan, China [1, 2] is responsible for causing the coronavirus disease termed COVID-19. SARS-CoV-2 spreads *via* small droplets and by fomites. The average number of people that one person with SARS-CoV-2 infects ranges from 2.0 to 2.5 persons [3]. Commonly reported symptoms of COVID-19 include cough, fever, shortness of breath, fatigue, initial nausea/vomiting, loss of smell, and taste [2, 4–6]. Approximately 20% of the infected cases progress to viral pneumonia, cytokine storm, and multi-organ failure [6]. As of June 25, 2020, there were 9,457,902 COVID-19 cases and 483,247 deaths reported worldwide according to the COVID-19 Dashboard by the Center for Systems Science and Engineering (CSSE) at Johns Hopkins University [7]. In the United States (US) alone, there were 2,381,369 COVID-19 cases (>30% of global cases) and 121,979 deaths with 28,567,355 reverse transcription-polymerase chain reaction (qRT-PCR) diagnostic tests [7]. With no vaccine available for SARS-CoV-2, implementation of wide-spread testing and contact tracing is critical to limit the transmission of the virus and allow gradual re-opening of the economy. However, shortages in supply chain, regulatory requirements and requirements for testing performed only by healthcare workers have restricted access to diagnostic testing and put further strains on the limited supply of personal protective equipment.

In response to the urgent public health need for accurate, effective, low-cost, and scalable COVID-19 testing technology, the Mass General Brigham Center for COVID Innovation (MGBCCI) launched a Diagnostics Direct-To-Consumer (DTC) working group to evaluate, identify and deliver viable testing solutions with potential for at-home usage that could eventually be implemented within the Mass General Brigham healthcare system and the broader local and global community. The US Food and Drug Administration (FDA) defines DTC as diagnostics that are “marketed directly to consumers without the involvement of a health care provider” [8]. The consumer collects a specimen (*e*.*g*., blood from a finger prick, saliva, or urine), runs the test, and receives the result within minutes. An example of a low complexity, at-home, DTC test for an infectious disease is the OraQuick^®^ in-home HIV antibody test [9]. Our objective in this investigation was to identify evidence-based and clinically informed specification criteria for DTC tests for implementation in Massachusetts to provide criteria that may be applied nationwide. This comparative evaluation of antigen and serology-based SARS-CoV-2 immunoassays (IA), and contact tracing solutions may accelerate innovative strategies that are low-cost and at-home products. Specifically, we aimed to establish a comprehensive list of currently available COVID-19 rapid test products with the potential to be administered in a DTC setting; and develop a framework for filtering and scoring to identify top candidates for evaluation in the MGBCCI Diagnostics Accelerator (DA).

To this end, we defined specifications required for viable rapid antigen and antibody tests, performed a deep horizon scan of products with potential for DTC at-home usage, and applied a heuristic scoring algorithm using test performance metrics along with the company, manufacturing, and supply-chain data. Due to the limited availability of molecular diagnostic antigen-based tests for COVID-19 with DTC at-home potential at the outset of our horizon scan, we focused primarily on serology-based tests. Companies having product information that meets the predefined specifications were engaged. Arrangements were made to evaluate those specification claims in the MGBCCI Diagnostics Accelerator for the following products: BodySphere, BioHit, BTNX, Edinburgh Genetics, Hangzhou Testsealabs, Intec Products, LumiQuick Diagnostics, Livzon Diagnostics, Meridian Biosciences, Ozo Life, Phamatech, RayBiotech, U2U Systems, and Vivacheck (see Table 1).

**Table 1.** Rapid serology-based lateral flow assays acquired for evaluation.

| **Company** | **Product** | **Status** | **Acquisition Time (days)** |
| --- | --- | --- | --- |
| BioHit Healthcare Co., Ltd. | SARS-CoV-2 IgM/IgG Antibody Test Kit | Queued for evaluation | 4 |
| BTNX Inc. | Rapid COVID-19 IgG/IgM Test Cassette | Queued for evaluation | 35 |
| Edinburgh Genetics Ltd. | COVID-19 Colloidal Gold IgG/IgM Testing Kit | Queued for evaluation | 4 |
| Genobio Pharmaceutical Co., Ltd.<br>(Subsidiary of Era Biology Group) | COVID-19 IgG/IgM Anti-body Lateral Flow | In Transit |  |
| InTec Products Inc. | Rapid SARS-CoV-2 Anti-body (IgM/IgG) Test | Under evaluation | 5 |
| LumiQuick Diagnostics Inc. | QuickProfile 2019-nCoV IgG/IgM Combo Test | Under evaluation | 2 |
| Meridian Biosciences | QuikPac II™ COVID-19 IgG and IgM Test | Queued for evaluation | 1 |
| Ozo Life Ltd. | Diamond SARS-CoV-2 IgM/IgG Rapid Test Kit | Queued for evaluation | 23 |
| Phamatech<br>(Distributor for Nanjing Vazyme) | COVID-19 Rapid Test IgG/IgM | Queued for evaluation | 6 |
| RayBiotech | Coronavirus (COVID-19) IgM/IgG Rapid Test Kit | Queued for evaluation | 1 |
| Shanghai Kehua Bio-Engineering | SARS-CoV-2 IgM/IgG Antibody (Colloidal Gold) | Queued for evaluation | 8 |
| Sure Bio-Tech Co., Ltd. | Coronavirus (COVID-19) Rapid Test | Queued for evaluation | 4 |
| Testsealabs Biotechnology Co., Ltd. | One Step SARS-CoV2 (COVID-19) IgG/IgM Test | Queued for evaluation | 43 |
| U2U Health<br>(Distributor for Cell-ID Pte Ltd.) | 2019-nCoV IgG/IgM Kit (Colloidal Gold-Based) | Queued for evaluation | 2 |
| VivaCheck Biotech Co., Ltd. | COVID-19 IgM/IgG Rapid Test | Queued for evaluation | 23 |
| Zhuhai Livzon Diagnostics Inc. | Diagnostic Kit for IgM/IgG Antibody to SARS-CoV-2 | Queued for evaluation | 6 |

In addition to molecular and serological tests, we identified digital solutions that may be coupled with biological tests to potentially offer a comprehensive solution and thereby mitigate the impact of the pandemic. Specifically, we examined several available electronic-based contact tracing solutions. We investigated the possibility of combining their use with the use of symptom self-reports collected with electronic patient reported outcome (ePRO) platforms and physiological data gathered with wearable sensors. These technologies are part of a broader category of devices referred to as mobile health (mHealth) technology. The WHO defines mHealth as “the use of mobile wireless technologies for public health” [3]. Over the past decades, we have witnessed tremendous growth in the use of mHealth technologies for the clinical management of individuals with conditions related to a broad spectrum of metabolic, cardiopulmonary and gastrointestinal diseases, neurological disorders, mental health, and environmental exposures [10, 11]. mHealth technology has many potential applications in the context of the COVID-19 pandemic. Herein we considered technologies that could augment if not replace manual contact tracing, and that could be combined with rapid antigen and antibody tests to improve the ability of health officials to detect and isolate COVID-19 cases. ePRO platforms can be used to track symptoms, which in turn can be used to locate geographical areas affected by outbreaks. They can be used to determine if individuals should be tested for COVID-19 based on reporting symptoms such as fever, dry cough, fatigue, sputum production, and shortness of breath or dyspnea, that have been associated with COVID-19 [2–4]. ePRO platforms can also be used to capture non-specific symptoms such as sore throat, myalgia or arthralgia, chills, headache, gastrointestinal issues, nasal congestion, hemoptysis, and conjunctival congestion that can be observed in COVID-19 patients [2–4]. Anosmia and ageusia have also been reported as possible COVID-19 symptoms [12].

Furthermore, cutaneous manifestations have been recently associated with COVID-19 [13]. Wearable sensors (Figure 1) can be used to augment ePRO measures by providing objective physiological data. It has been argued that data that can be collected using wearable sensors, such as measures of heart rate (HR) at rest [14] and cough frequency [15], could be associated with COVID-19 and utilized to identify individuals who should be tested because they show early signs of the disease. Finally, mHealth technology can be used to trace people and assess the likelihood of infection *via* contacts with other individuals who tested positive for COVID-19. The use of “big data” has the potential to limit the spread of the disease by strengthening disease surveillance, monitoring for health decline and adverse events, and inform on transmission *via* tracking the population [16]. When this is done in the community, the enabling technology is referred to as “community tracing.”

**Figure 1.**
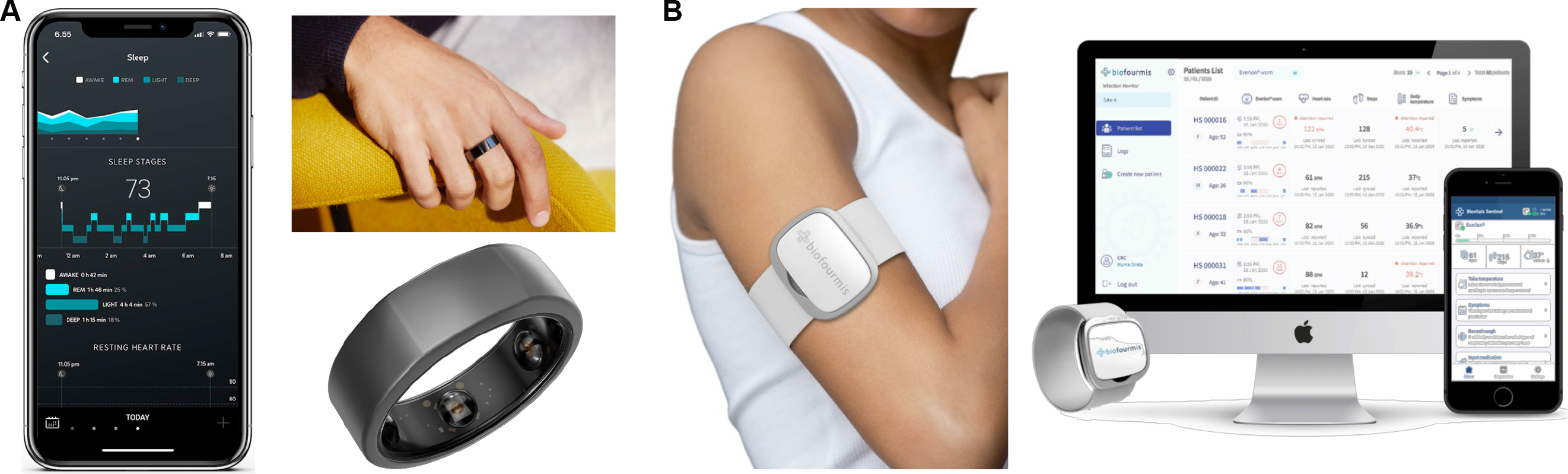
Examples of wearable technology. A) the Oura Smart Ring sensor and an image of the app utilized to collect and display physiological data collected using the sensor. B) the Everion system by Biofourmis. The system is used to collect data such as heart rate, respiratory rate, and oxygen saturation. Images reproduced with permission by the manufacturers.

To examine the above mentioned technologies, we assembled a Working Group of nearly 60 individuals with expertise in mHealth technology (electronics, computer science, signal processing, and machine learning as well as in clinical areas relevant to COVID-19) to survey commercially available systems, establish a framework to evaluate existing technologies, and identify gaps and future work to address them.

## Methods

### Specifications for Molecular and Serological Tests

#### Literature and Internet Search Strategy

US FDA, Foundation for Innovative New Diagnostics (FIND), The Center for Systems Science and Engineering (CSSE) at Johns Hopkins University (JHU), Pubmed, bioRxiv, medRxiv, and news articles as well as PubMed database for articles on “SARS-CoV-2” and “diagnostics” were searched. The Medical Subject Headings search terms including “COVID-19”, “SARS-CoV-2”, “coronavirus”, “antigen testing”, “PCR”, “antibody testing”, and “IgG” were used.

#### Survey

Expert interviews with two infectious disease physicians and one pathologist testing point-of-care assays at Brigham and Women’s Hospital (BWH) were conducted using Zoom or phone. Also, a short, online survey to identify key specification metrics was developed. The survey was circulated to clinicians and epidemiologists working in infectious diseases, emergency medicine, intensive care, and other clinicians and researchers responding to the COVID-19 pandemic. The respondents were affiliated with (BWH), Massachusetts General Hospital (MGH), as well as external colleagues of the sub-group. Survey questions included:

1. You have received this survey because you are involved in response to the COVID-9 pandemic. What is your primary role?
2. What is your primary affiliation?
3. Is there an unmet clinical need where serological tests (blood from a finger prick, saliva, etc.) for SARS-CoV-2 antibodies can help?
4. At what level of specificity, sensitivity, PPV, NPV would a DTC serology-based antibody test for immunity to COVID-19 be useful for a “return to society” scenario?
5. Are you collecting data or can recommend a colleague or a company that is collecting data on the relationship between IgG positivity and transmission (please include your email if we can contact you about data)?
6. How likely is it that you would recommend an at-home/DTC serology-based antibody test to a patient?
7. In your own words, what do you want from a DTC serological test for the SARS-CoV-2 antibody?
8. In your own words, in the DTC setting for diagnostic testing, what is the value of a viral antigen or molecular test?
  a. In the DTC setting for diagnostic testing for the viral antigen, what type of self-collection method (nasal swab, oral swab, saliva, *etc*.) would be most accurate?
  b. What level of sensitivity, specificity, accuracy is needed for a rapid, viral antigen test to be useful to lift the “stay-at-home” scenario?
9. Is there a DTC platform for COVID-19 (or prior infectious disease outbreaks) that you have evaluated or recommend (please include your email address if we can contact you about your expertise with DTCs)?

85 physicians or epidemiologists responded to the survey (see Figure 2). The majority of survey respondents saw value in DTC antibody testing, especially for assessing the prevalence of the SARS-CoV-2 exposure in the population, would recommend DTC tests, and felt antibody tests would inform a return-to-work scenario.

**Figure 2.**
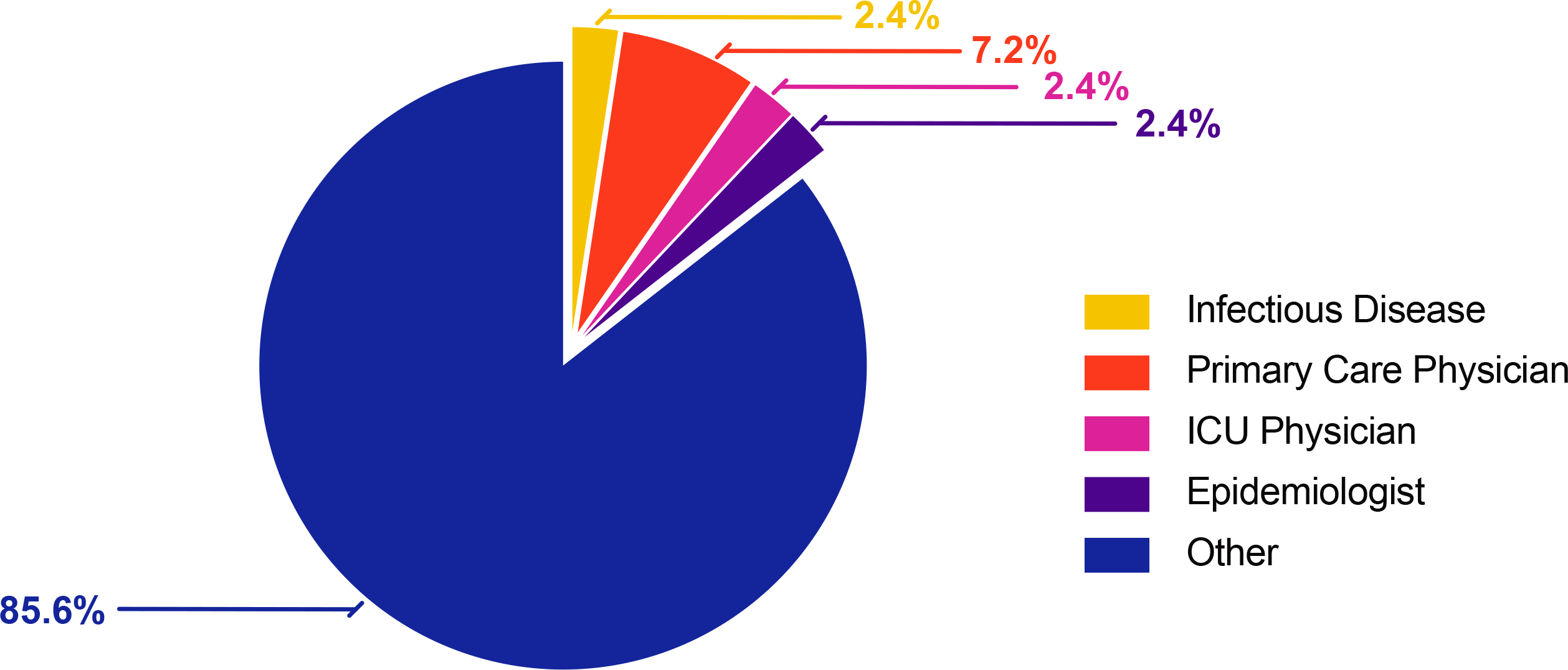
Survey responders. The graph shows the percentage of responders’ medical specialty. Other category includes several dermatologists, dermatologist & immunologist, skin cancer surgeon, several pathologists, allergist/immunologist, neurologists, division chief of neurology, ICU nurse practitioner, OB epidemiologist, radiologist, specialty physician, medical oncologist, anesthesiologist, visiting scholar, several research fellow or research scientist, pharmacist, leadership, management of operations, hematologist.

#### Specification Criteria

Figure 3 illustrates the iterative application of the specification criteria to systematically review and filter over 300 tests identified on the FDA, FIND, and JHU websites.

**Figure 3.**
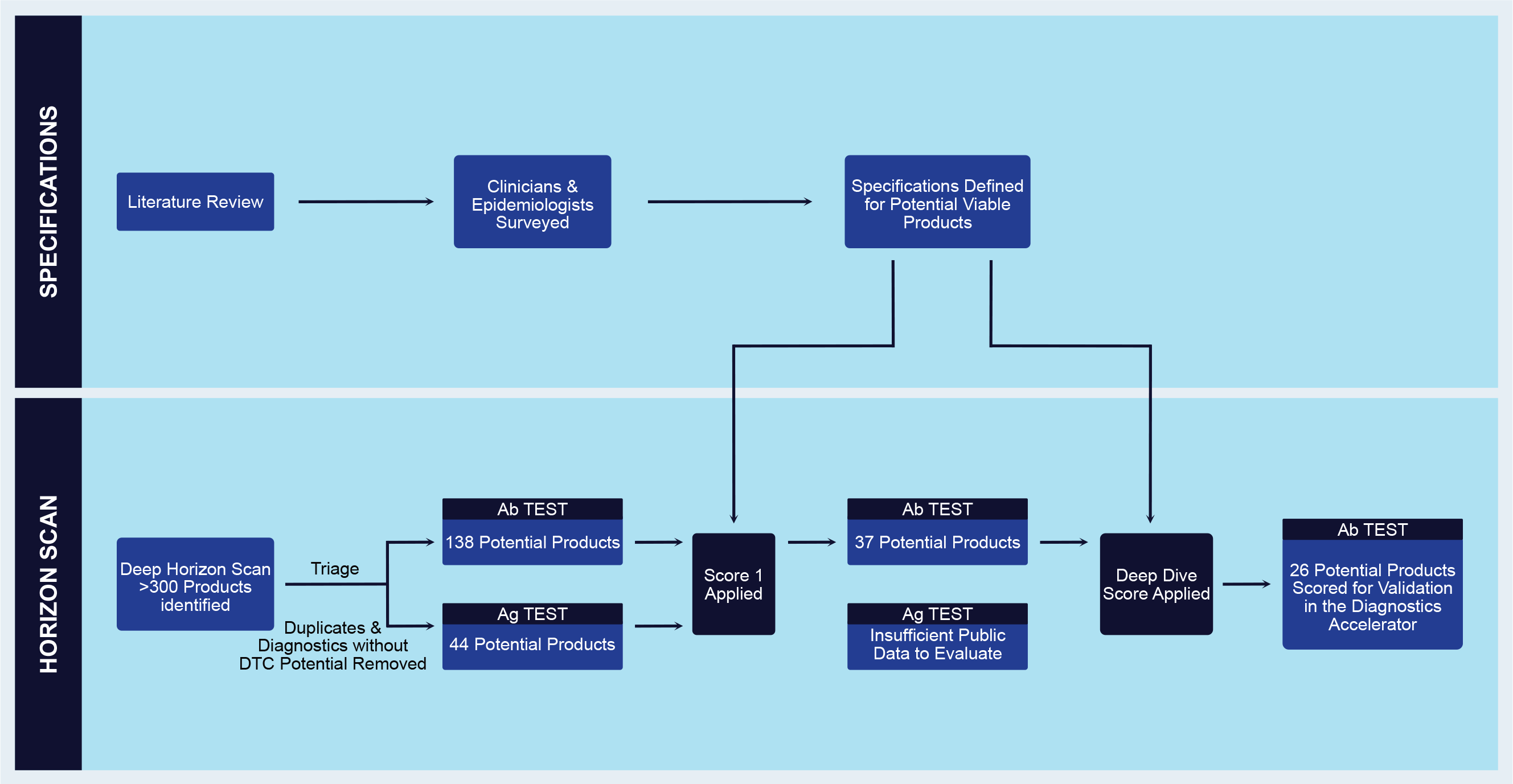
Horizon scan work flow. The list of antigen and antibody tests for SARS-CoV-2 was filtered based on data from survey to clinicians, data extracted from company websites and reach out to obtain key data on performance metrics, validation study, and scalability.

#### Regulatory Status

Regulatory status for each test, including FDA clearance, approval in and outside the U.S., and FDA EUA status was evaluated.

#### Performance Metrics

Sensitivity is the percent of individuals with the disease correctly classified by the test as having the disease. For antigen testing, we considered highly sensitive tests (95-99%). For antibody testing, we considered sensitivity over 95%. Specificity is the percentage of individuals without the disease correctly classified by the test as being disease-free. We gave preference to products with a specificity of 99% for antibody testing.

#### Sample Source

Sampling sources, including nasopharyngeal swab, nasal swab, and saliva, were considered in the scoring for antigen testing. For antibody tests, only blood from a finger-prick was recognized as a sample source in the scoring algorithm. However, new tests still in development are detecting antibodies in saliva, so this parameter may need to be adjusted in the future to encompass all products with DTC potential.

#### Biological Target

SARS-CoV-2 has four structural proteins: spike [S], membrane [M], envelope [E], and nucleocapsid [N] proteins. SARS-CoV-2 binds with a high affinity to the Spike protein receptor-binding domain (RBD) angiotensin-converting enzyme 2 (ACE2) and uses ACE2 as an entry receptor to invade the host cells. We hypothesized that anti-IgG to the Spike protein would be more specific and weighted this parameter according to the scheme described in the Horizon Scan section.

#### Antibody Assay and Detection Method

The common types of immunoassays (IA) are rapid lateral flow IA (LFIA), manual ELISA, and automated chemiluminescent IA (CLIA) [17]. LFIA provides a rapid, immunochromatographic reaction with a subsequent visual colorimetric read-out within 0-30 minutes after initiation of the test. The principle relies on antigen-antibody interactions and capillary action of analytes moving through a polymeric membrane, often divided into distinct sections. LFIA’s contain one control line to ensure the functionality of the test and one or more test lines for antibody detection. Upon the detection of monoclonal antibodies in the serum, a visual read-out is obtained in the form of a distinct, colored line. The test is disposable and can be stored at room temperature [18]. These tests are packaged in easily used cassettes and are affordable and easy to use. Once the appropriate supply chain components are in place, the tests can be manufactured at scale to meet the needs of the global pandemic. Such parameters were key specification criteria critical to the next phase of the product evaluation.

### Horizon Scan

We studied the literature with the goal of comprehensive coverage of promising tests both in-use with potential DTC application and those in the concept or developmental phase. Lists published by FIND, FDA, Johns Hopkins University, and RTT News served as first-pass sources, and additional tests were added to the long-list based on the broad online searches. The preliminary scan was completed over two weeks during April 2020, and results reflect the information available at that time. The following attributes were collected for each test in the preliminary scan:

i. Name of organization
ii. Type of organization (Corporation, Academic, other)
iii. Organization location
iv. Name of the product and product number, if available
v. Type of test (antibody, antigen, other)
vi. Whether the test was currently in-use (Y/N), and if in-use, where
vii. FDA approval status
viii. Approval status with international regulatory bodies
ix. Current application type (Point-of-care, Lab-based, Research-only, DTC)

Following the preliminary scan, the Horizon Scan team worked together with the Specifications team to establish a methodology for scoring candidate tests. As the methodology development was partially dependent on the results of the scan, we have elaborated on scoring methods in the Results section. The scoring methodology was designed to first filter down the long-list to a shorter list of targets for deep-dive research, and then to focus the deep-dive research on a set of parameters that would be used to calculate a first relative score for each test (SCORE 1). The final algorithm for SCORE 1 was the following:

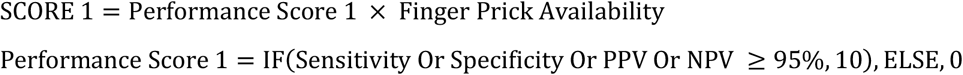

The Finger Prick Availability score was binary, with a score of 1 if the test could be administered using a finger prick. After the top candidates had been assigned a value for SCORE 1, we determined a set of parameters and weight values for a second, and more stringent score (SCORE 2), designed to establish the priority of each candidate test for further performance validation work. Representatives for top candidate tests were contacted during the SCORE 1 and SCORE 2 research periods and asked to provide performance and other data not attainable through public sources of information. The final algorithm for SCORE 2 was the following:

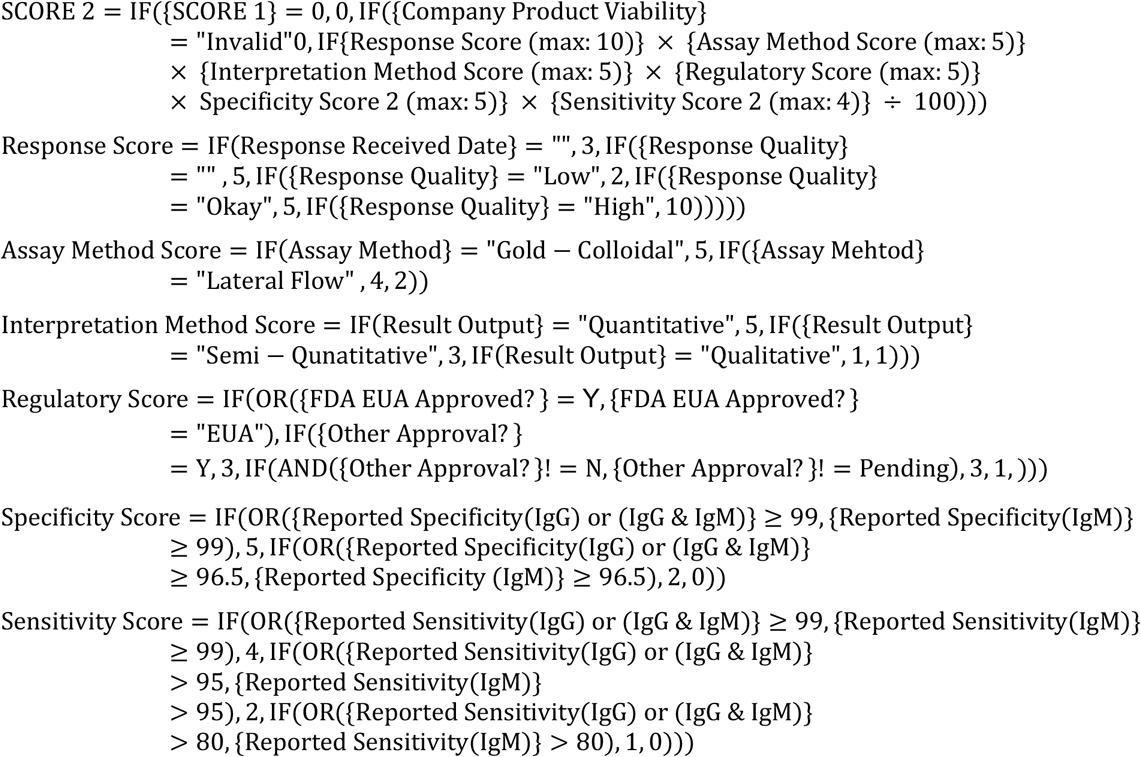

Upon assignment of SCORE 2, findings were formatted in a responsive tabular format (Airtable) designed to be embedded in a website for reference by the public. This embedded table was linked to the working table used by the Horizon Scan team, so that any updates made by the Horizon Scan team to the available information on each candidate test, or addition of new tests, will be automatically reflected on the public website.

### Approach to Identify Suitable mHealth Technologies

Commercially available electronic patient-reported outcome (ePRO) platforms were examined to determine their clinical utility and potential implementation during the COVID-19 pandemic. Figure 4 shows an example of the ePRO platform for COVID-19 self-reports [19]. Twelve systems were analyzed. Evaluation of their specifications was performed by considering the versatility of the solution type, availability of the platform in different operating systems, the functionality of the user interface, and regulatory compliance. Clinical utility and customization of the medical questionnaires incorporated into the different ePRO solutions were determined by analyzing the extent and relevance of the included signs and symptoms. Parameters, including severity, localization, duration, and progression of signs and symptoms, were considered to evaluate the quality of the clinical assessment. Features such as direct communication between patients and clinicians, geo-localization, and telemedicine integration were assessed to determine their availability and interconnectivity within the same system and other services. Integration with external platforms such as wearable technologies and electronic medical record (EMR) systems was investigated and documented, and the ability and performance of ePRO solutions to collect, transfer, visualize and analyze data were evaluated.

**Figure 4.**
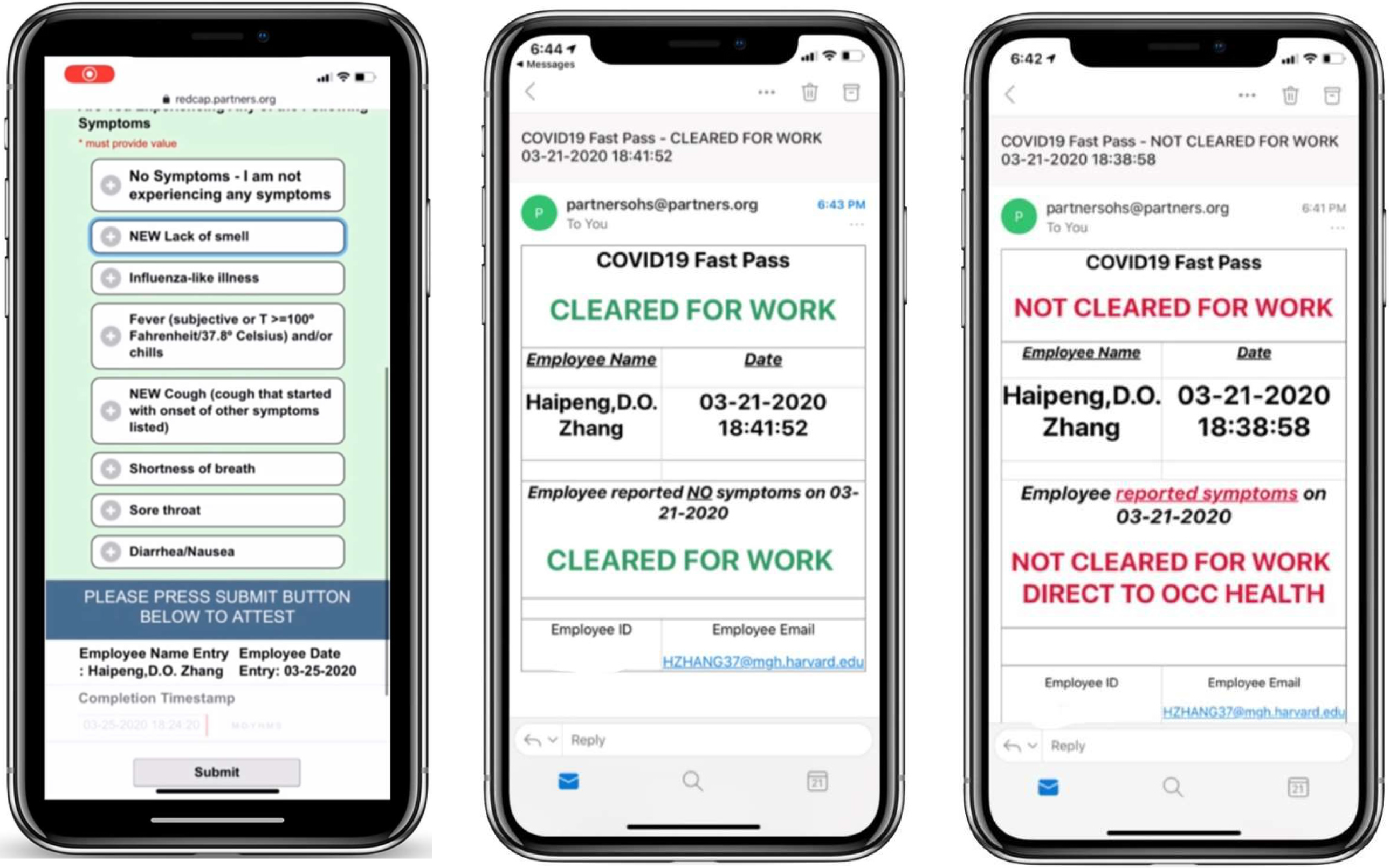
Screenshot of the platform currently utilized by MGB to collect self-reports of COVID-19 symptoms. Image reproduced with permission from Ref. [19].

A survey of existing sensing technologies was conducted to investigate the feasibility of monitoring physiological and physical variables that are relevant to the symptoms of COVID-19. General attributes typically used to assess mHealth technologies, as well as specific characteristics pertinent to achieving early detection of infection in individuals who are either pre-symptomatic or asymptomatic, were considered. Also, characteristics that could be continuously monitored using mHealth solutions versus those that could be reliably captured using ePRO solutions were identified. The clinical features deemed appropriate to be monitored using mHealth technologies included fever, cough, fatigue, dyspnea, hypoxemia, and high systolic blood pressure. In contrast, it was deemed more appropriate to use ePRO solutions to monitor the production of sputum and hemoptysis, and to detect when patients have muscle pain, headache, diarrhea, and chills. Primary physiological parameters that were deemed relevant to or indicative of the above-mentioned clinical variables and monitored using existing on-and/or off-body sensing technologies include body temperature (fever), number and character of coughs (cough), motion or activity level (fatigue, falls), respiration rate (RR; dyspnea), oxygen saturation level (SpO2; hypoxemia), and blood pressure (BP) (*i*.*e*., high systolic BP). Also, heart rate (HR), HR variability, and electrocardiogram (ECG) data were also considered as physiological data of potential interest in the context of COVID-19. This is mainly because there is increasing recognition of a primary effect on cardiac function during late incubation, early prodromal, illness, decline, and convalescence stages of the disease.

To evaluate solutions for contact tracing in the community, we identified characteristics enabling the assessment of their suitability to detect COVID-19 cases and their contacts. Based on a preliminary search of contact tracing solutions, we decided to include online surveys and data aggregator solutions that could directly or indirectly support smartphone-based contact tracing. Solutions were evaluated based on publicly available information. When possible, developers were interviewed to gather the following information. How is the solution delivered to the end-user? Is it a Smartphone App, an Online Survey, or a Data Aggregator? Which operating systems (iOS, Android) are supported by the solution? This was limited to smartphone app solutions. How is the solution deployed? Is it a solution that can be used on its own, or is it a solution that is implemented as a part of another system? What type of data does the solution collect? The following list was considered: Age, Gender, Location, Proximity, Phone Number, IP Address, Contact Duration, Symptoms, Zip Code, Travel Mode, Trip Purpose, COVID Diagnosis, Travel History, Medical Conditions, Temperature, COVID Contact, Quarantine Status, Race, Ethnicity, Household Info, Health Insurance Coverage, Email Address, Case Statistics by Location, Age Group, Occupation, Smoking Status, Medications, Flu Vaccine Status, Population Mobility, Behavioral Insights, Contact Distance. Is the solution open source or proprietary? Does the solution preserve the privacy of the end-user? Does the solution track the location of the end-user? What is the privacy policy of the solution? What is the availability status of a solution? Is the solution currently available? Is it under development? Is the solution at a concept stage and development has not started yet? Where can the solution be deployed (***i***.***e***., in which geographical areas)?

## Results

### Horizon Scan and Scoring Algorithms

The preliminary scan resulted in a long-list >300 potential products, of which >120 were deemed duplicate or too incomplete for further research (Figure 3). We then progressed to the scoring phase, with additional updates made to the long-list regularly. The Australian Government Department of Health (ARTG) list of COVID-19 test kits for legal supply in Australia resulted in several additions.

Next, duplicate products were removed from the curated list, and the 178 different tests identified from the FDA and FIND websites were separated into 44 antigen and 138 antibody tests. For viral antigen testing, we determined regulatory status (*e*.*g*., FDA cleared or authorized, approved outside the U.S.), performance (*e*.*g*., sensitivity), sampling method (*e*.*g*., saliva, blood), supply chain availability, manufacturing capability, and scalability as crucial specification metrics (Table 2). We identified regulatory status, performance (*e*.*g*., specificity and sensitivity), and biological target (SARS-CoV-2 spike (S) protein or nucleoside (N) protein), supply chain availability, manufacturing capability, and scalability as key specification metrics for DTC antibody testing (Table 3). An additional metric considered was cost.

**Table 2.** Specifications for DTC antigen tests.

| Parameter | Approval | Survey Resonders Preference |  | Survey Resonders Preference |
| --- | --- | --- | --- | --- |
| Regulatory | FDA Approval | Preferred |  |  |
|  | Approved outside of the US |  |  |  |
|  | Awaiting FDA |  |  |  |
|  | Unknown |  |  |  |
| Performance | **Sensitivity<sup>a</sup>** |  | **Specificity<sup>a</sup>** |  |
|  | >99% |  | >99% | Preferred |
|  | 95-99% | Preferred | 95-99% |  |
|  | <95% |  | <95% |  |
| Sample | **Source** |  |  |  |
|  | Saliva | Preferred |  |  |
|  | Proximal nose |  |  |  |
|  | Nasopharyngeal |  |  |  |

<sup>a</sup> IgG or IgG+IgM

**Table 3.**
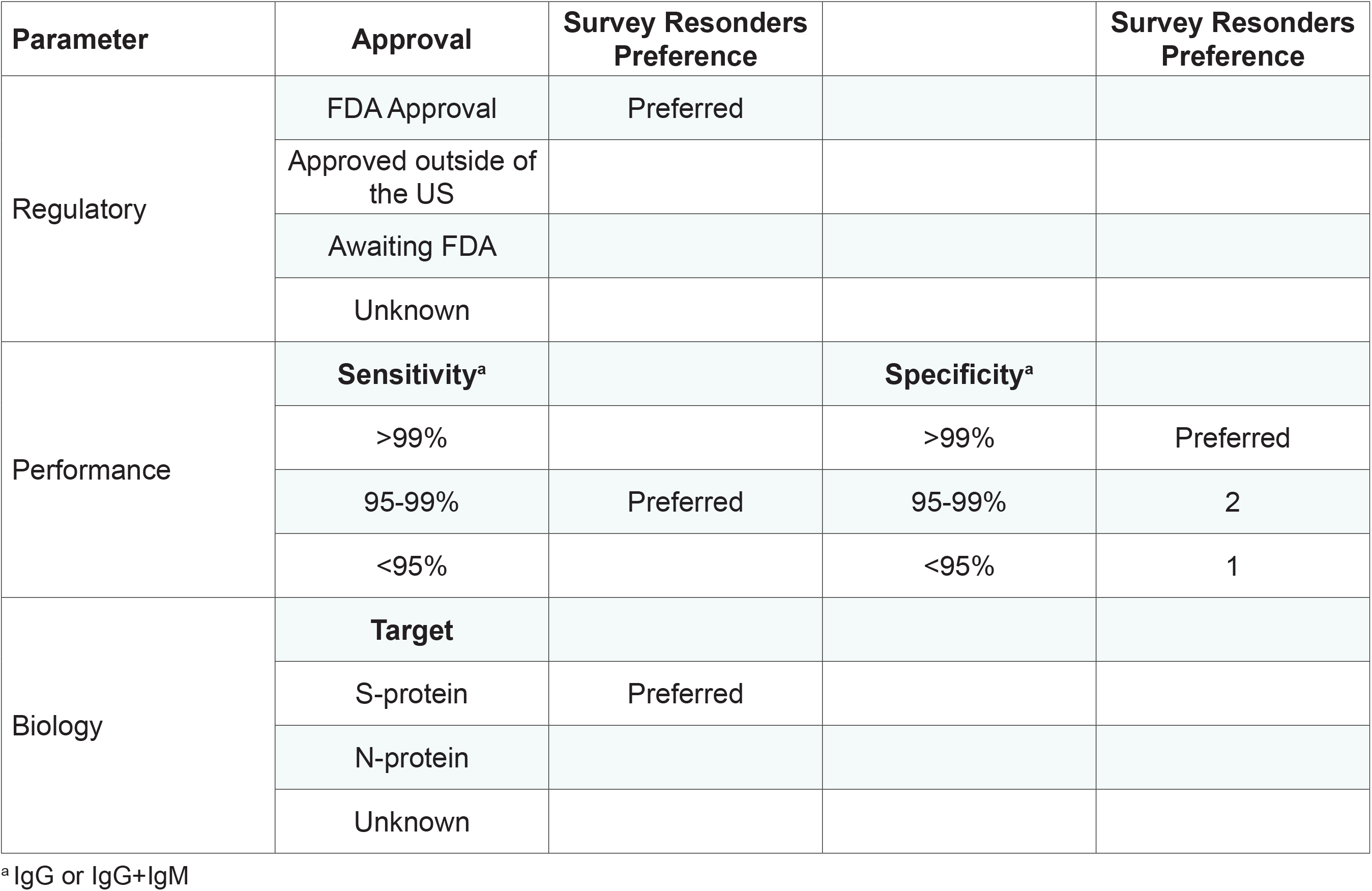
Specifications for DTC antibody tests.

| Parameter | Approval | Survey Resonders Preference |  | Survey Resonders Preference |
| --- | --- | --- | --- | --- |
| Regulatory | FDA Approval | Preferred |  |  |
|  | Approved outside of the US |  |  |  |
|  | Awaiting FDA |  |  |  |
|  | Unknown |  |  |  |
| Performance | **Sensitivity<sup>a</sup>** |  | **Specificity<sup>a</sup>** |  |
|  | >99% |  | >99% | Preferred |
|  | 95-99% | Preferred | 95-99% | 2 |
|  | <95% |  | <95% | 1 |
| Biology | **Target** |  |  |  |
|  | S-protein | Preferred |  |  |
|  | N-protein |  |  |  |
|  | Unknown |  |  |  |

<sup>a</sup> IgG or IgG+IgM

The primary filter utilized two binary attributes to restrict SCORE 1 research to the top candidates (Tables 4 and 5). These attributes were (i) feasibility of use outside of the hospital setting and (ii) whether the test was currently in-use or had any significant regulatory activity. If the information on these attributes was not easily obtained through searching, the test was also removed from consideration. During the research process, it became clear that performance data were not readily available for many of the antigen test candidates. This was likely due to the early stage of development of these candidates. We then decided to pursue only antibody tests for further evaluation, with updates to the antigen test attributes planned to run in parallel. The parameter values and weights of SCORE 1 were also adjusted during this period, due to changes in information available on the clinical value of specificity, sensitivity, and biological target in COVID-19 antigen tests (for final weights, see Table 4 and Table 5). Upon application of the primary filter, there were a total of 65 candidates remaining (17 antigen tests and 73 antibody tests). Upon application of SCORE 1, 37 antibody tests were remaining with a score higher than zero.

**Table 4.** Specifications for DTC antigen tests based on updated score 1.

| Parameter | Options | Option Weight |
| --- | --- | --- |
| Performance | Sensitivity | >95% |
|  | Specificity | >95% |
| Sample Source | Saliva | 5 |
|  | Proximal Nose | 3 |
|  | Pharynx | 1 |

SCORE 1 = Performance Score \* Sample Source
Performance IF (Sensitivity OR Specificity OR Positive OR Negative predictive values $\geq 95\%$ ), 10, else, 0
Finger Prick Availability See “Option Weight” above

**Table 5.** Specifications for DTC antibody tests based on updated score 1.

| Parameter | Options | Option Weight |
| --- | --- | --- |
| Performance | Sensitivity | >80%=1 |
|  | Specificity | >99%=5, >96.5%=1, <96.5%=0 |
| Finger Prick Availability | Yes | 1 |
|  | No | 0 |

SCORE 1 = Performance Score \* Finger Prick Availability
Performance IF (Sensitivity OR Specificity OR Positive OR Negative predictive values $\geq 95\%$ ), 10, else, 0
Finger Prick Availability See “Option Weight” above

After the top candidates had been assigned a value for SCORE 1, we determined a set of parameters and weight values for a second, and more stringent score (SCORE 2), designed to establish the priority of each candidate test for further performance validation work (see Table 6). We then conducted a deep-dive to extract data on biological target, assay method, detection method, and other vital parameters. We searched for publicly available information on the best contact person, assay type, and interpretation method for each test. Then, a MGBCCI email address was used to solicit additional information from each candidate organization, including a link to a survey to fill out SCORE 2 parameters (see Table 7). After 24 hours, we made follow-up phone calls to unresponsive organizations. Out of 37 individual outreach attempts, we received a response from 26 of them by May 22. Responses were further categorized by level of detail and perceived interest in pursuing additional performance testing. A deep dive was then applied to all 37 remaining candidates. As the current SCORE 2 value for each test frequently updates based on the inclusion of additional information, please see the results webpage (https://covidinnovation.partners.org/diagnostics-direct-to-consumer/) for the final output.

**Table 6.** SCORE 2 specifications for DTC antibody tests that passed score 1>0 only.

| Parameter | Options | Option Weight |
| --- | --- | --- |
| Performance | Sensitivity | >99%=4, >95-99%=2, >80%=1 |
|  | Specificity | >99%=5, >96.5-99%=2, ≤96.5%=0 |
|  | PPV | NA |
|  | NPV | NA |
| Regulatory | FDA Approval | 5 |
|  | Approved outside of the US | 4 |
|  | Awaiting FDA, "in-development" | 1 |
| Assay Method | Gold-Colloidal | 5 |
|  | Lateral Flow | 4 |
| Interpretation | Quantitative | 5 |
|  | Semi-quantitative | 3 |
|  | Qualitative | 1 |
| Responsiveness | Response quality "HIGH" | 10 |
|  | Response quality "OK" | 5 |
|  | Response quality "LOW" | 2 |
|  | NO Response yet | 3 |

**Table 7.**
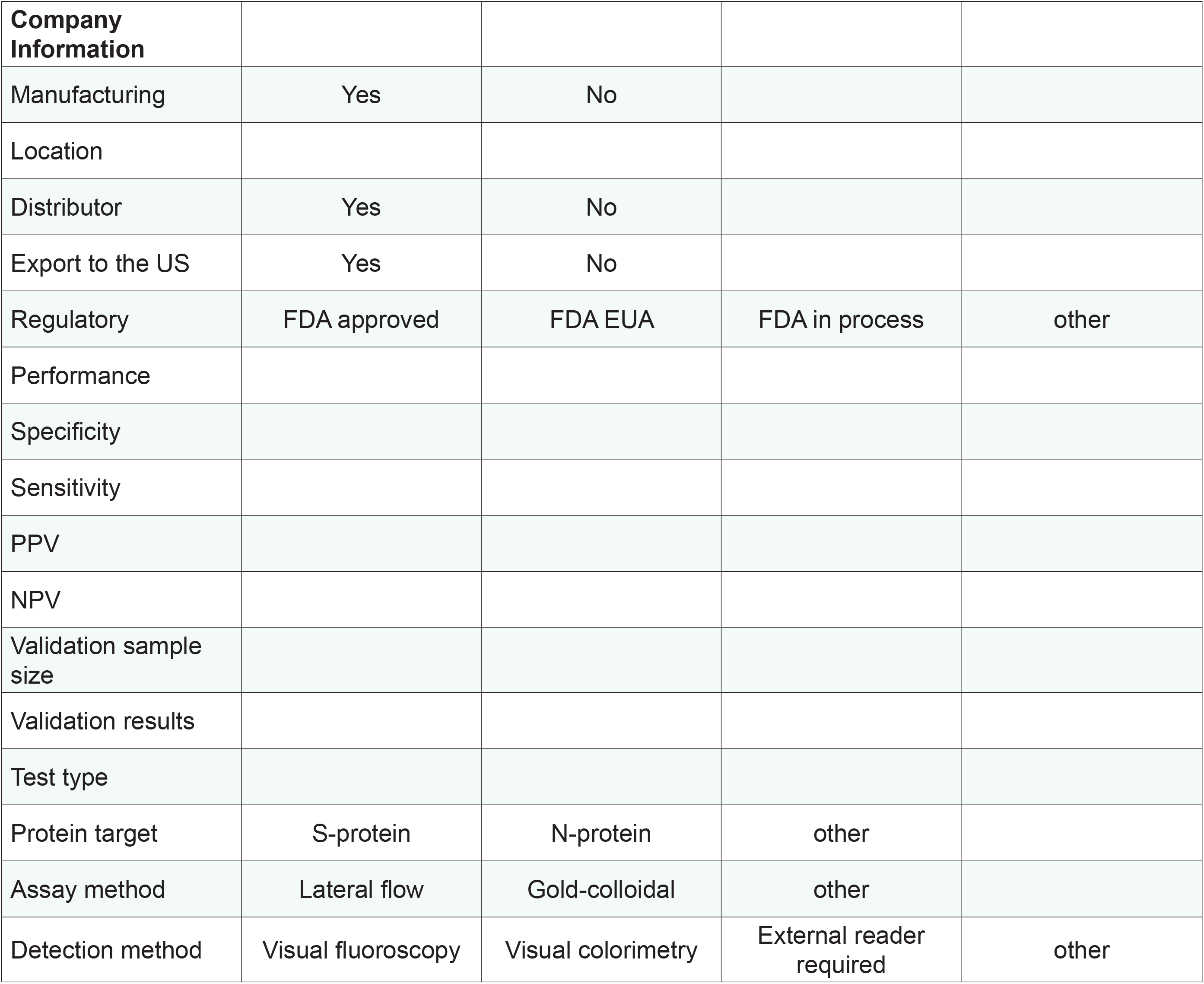
Informational fields from deep-dive survey.

### Suitable mHealth Technologies

Numerous commercial ePRO solutions have implemented specialized COVID-19 modules to assess related symptoms in the form of clinical questionnaires. Several of these platforms enable the stratification of subjects according to their risk of infection. The analyzed solutions use the following main factors as part of their clinical algorithms to classify patients: demographics (age and gender), exposure to potentially infected patients, signs, symptoms, comorbidities, and other relevant conditions [20]. Symptoms utilized by most ePRO solutions include common COVID-19 symptoms (*i*.*e*., fever, tiredness, and dry cough) as well as less common symptoms such as diarrhea, aches, and pain [21]. Most algorithms classify shortness of breath and respiratory distress as a severe symptom that should be promptly managed at a hospital [22]. Some comorbidities such as chronic obstructive pulmonary disease, obesity, coronary artery disease, hypertension, diabetes, cancer, and immunosuppression are known to increase the mortality of COVID-19 patients and are considered in the implementation of clinical algorithms [23]. Patient stratification is also performed by age because the disease causes higher mortality among older adults; estimated mortality rate as high as 5-20% had been reported for individuals 80-90 y/o or as low as 0.2% for patients 20 y/o and younger. Some algorithms take into consideration whether the subject is in a geographical area where COVID-19 is prevalent, thus increasing the risk of exposure and subsequent infection [24]. After the algorithm is implemented and the severity of the symptoms in conjunction with risk exposure, age, comorbidities, and other relevant conditions are assessed, the patient is managed with different clinical protocols. A low-risk patient with mild symptoms is encouraged to self-quarantine at home with follow-up questionnaires, telemedicine, or phone consultations. The presence of cardinal symptoms such as high fever, dry cough, and respiratory distress in combination with other risk factors will trigger urgent attention, connecting and directing the patient to emergency services and the closest healthcare facility.

We succeeded in identifying sensing technologies meeting criteria identified *via* group consensus and related to the following points: 1) technical/clinical validation and FDA 510(k) clearance or CE certification; 2) ease of use, and 3) integration with existing hospital systems to gather clinical data in the field. Specifically, we identified systems supporting the measurement of body temperature and respiratory rate that are more closely related to the prominent symptoms of COVID-19, but also provide a comprehensive set of measurements (*i*.*e*., SpO2, HR, and ECG) that could reflect subtle physiological changes during the pre-symptomatic phases of the disease. Additional factors related to the deployment of the technology were considered, such as level of user compliance, preference of wearing sensors during the day or at night, and durability (*e*.*g*., waterproof/showerproof). In the context of achieving early detection of exposure to COVID-19 patients, we identified solutions with potential for detecting early symptoms that are indicative of infection during the incubation period, such that infected individuals can self-isolate to avoid further spreading of the virus and receive immediate care to expedite the recovery process. The fundamental hypothesis that supports the feasibility of early detection of COVID-19 infection during the incubation period is that there is a gradual change in primary clinical symptoms (*e*.*g*., skin temperature, RR, SpO2, HR, ECG, *etc*.). This appears to be a reasonable hypothesis given that there is supporting evidence that analyzing physiological characteristics (*e*.*g*., ECG, hemodynamics, and temperature) in non-human primates exposed to different pathogens (*e*.*g*., Ebola) could yield early-detection of the infection [25]. After the study, we identified a total of ten example technologies that met the functional criteria for monitoring individuals in their home.

We identified 26 smartphone-based solutions that capture data for contact tracing. Of these, zero were open-source, seven implemented approaches for preserving privacy, and none of them required location tracking. At the time the group surveyed these technologies, eight apps were actively in use, three were under development, and five were at the concept stage. All of the active apps were available on both iOS and Android. Privacy policies were available for seven smartphone apps, but the scope and quality of information varied amongst the apps. Academic institutions or non-profit organizations were associated with three of the apps. TraceTogether (Government of Singapore), Aarogya Setu (Government of India), and Rakning C-19 (Iceland’s Civil Protection and Emergency Management team) are all government-developed apps that require users to provide their phone number so that manual contact tracers could follow up if needed. Aarogya Setu was available in 11 languages and had been installed more than 50 million times. Solutions like SafePaths, COVID Watch, and PACT are based on strict privacy-preserving protocols and only capture data related to proximity and/or location, other solutions capture information of a much broader range. We also identified a total of six online surveys designed to obtain information that could assist in contact tracing efforts. Five of the online surveys are currently active, and four of them are associated with academic institutions. Finally, we identified data aggregators that use data collected by existing location services (*e*.*g*., Google Maps) to provide population-level mobility insights and case statistics by location. Eight of these solutions are provided by companies that routinely capture location data of users for their business applications. The remaining solutions rely on these datasets but are maintained by either academic institutions, governments, or open-source projects. Corona Map (maintained by the Government of South Korea) and Corona Data Scraper (open source project) provide case statistics by location, whereas the rest provide population mobility statistics.

## Discussion

Our objective was to identify evidence-based and clinically-informed specification criteria for DTC tests for implementation in Massachusetts to provide criteria that may be applied nationwide. We aimed to (i) develop specifications, including performance criteria, scalability, and readiness, (ii) conduct horizon scanning for the most appropriate technologies and down-select to the most promising solutions, (iii) collaborate with the MGBCCI Diagnostics Accelerator and Validation working groups to ensure the viability of potential technologies, and lastly (iv) iterate with the Diagnostics Accelerator, Validation investigators, and the private sector to ensure timely delivery of DTC diagnostic solutions and assemble a clear plan for successful implementation.

### Identifying and Assessing Antigen and Serology-based Rapid Tests

Current BWH guidelines for HCWs to return-to-work are two negative PCR tests twenty-four hours apart. The DTC specifications data may accelerate innovative strategies for low-cost, at-home testing for HCWs. Early diagnosis of SARS-CoV-2 infection is one of the critical steps to control coronavirus transmission. Several diagnostic assays based on quantitative reverse transcriptase PCR (qRT-PCR) were rapidly developed after the discovery of SARS-CoV-2 [26]. qRT-PCR is the standard reference test for diagnosing SARS-CoV-2 with high sensitivity and accuracy in the early phase of infection. SARS-CoV-2 viral RNA load has been detected in nasal swabs and throat of coronavirus patients using qRT-PCR (Figure 5, grey box). The presence of the virus is undetectable by 14 days of post-symptom onset (Figure 5) [27, 28]. The presence of both SARS-CoV-2 IgM and IgG antibodies (Figure 5) are detectable as early as 3-4 days post-symptom onset [29–33]. IgM antibody peaks between 2-3 weeks and stay detectable after one-month post-symptom onset [30], while IgG antibody peaks after 5-6 weeks post-symptom onset [29, 31, 32]. Currently, there are no reports on the presence of these SARS-CoV-2 IgG antibody in the later phase of post-symptom onset. Based on the SARS-CoV-2 immune response timeline, the specification criteria were defined based on the use case for DTC testing for COVID-19 into two categories, based on days since symptom onset: 1) diagnosis with antigen testing and 2) seroconversion with antibody testing.

**Figure 5.**
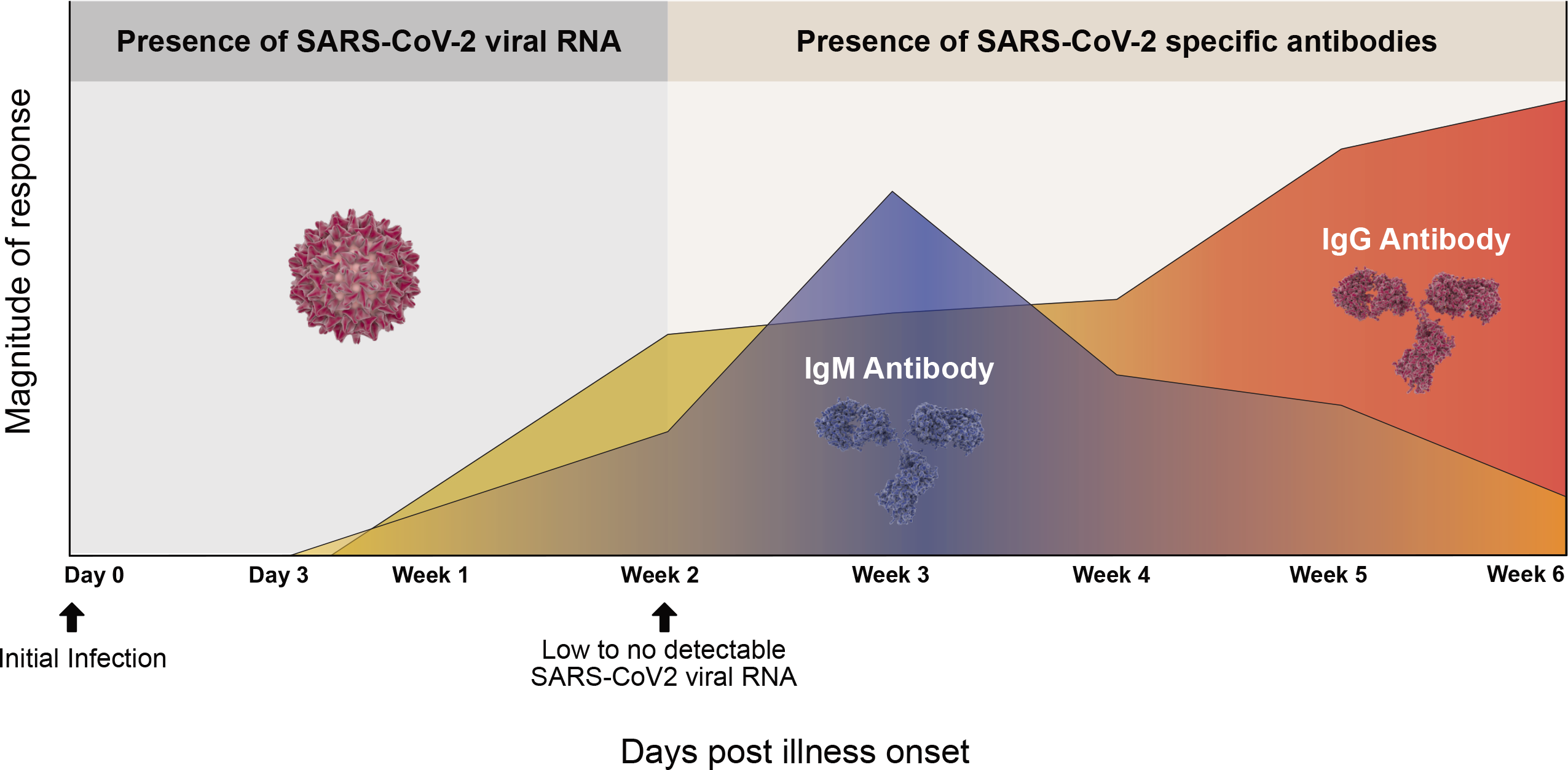
Schematic illustration of viral RNA and antibodies during the timecourse of SARS-CoV-2 infection. Data extracted from Refs. [27–33].

A COVID-19 antigen test detects the presence or absence of a SARS-CoV-2 viral antigen in a nasopharyngeal specimen obtained by using a flocked swab. An antigen is a structure within a coronavirus that triggers the immune system’s response to fight off the infection. Antigen tests that detect respiratory syncytial virus, influenza virus, HIV, or malaria antigens by immunoassay directly from specimens have been used for diagnosis. These tests have been commercially available for years, are of low complexity, and may provide results (<1 hour) at the point of care [34]. An antigen test is effective because it can take a few days for the immune system to build enough antibodies to be detected in a serology detection test; however, COVID antigens can be identified almost immediately after infection. So, in principle, the rapid antigen detection test can be an earlier source of information on whether someone has been infected with the virus. Current assays for other viruses, including influenza and respiratory syncytial virus, suffer from suboptimal sensitivity to rule out disease [35, 36]; the same challenge exists for SARS-CoV-2. Prototypes of antigen tests for other coronaviruses are still are under development and have not yet received regulatory approval [37, 38]. Monoclonal antibodies against the SARS-CoV-2 protein have been produced, and this might set the foundation for a future rapid antigen test [39]. An additional hurdle for the production of a rapid antigen test that can be used as a DTC product will be in its ability to retain the necessary sensitivity when using saliva or blood from a finger prick, as both sources are known to contain significantly less viral load than pharynx samples. One other issue is the potential for cross reactivity with proteins from other coronaviruses, such as those that cause some forms of the common cold, which would lead to false positive results.

SARS-CoV-2 utilizes the receptor-binding domain (RBD) of its Spike protein to bind with high affinity to the host cell’s transmembrane protein, angiotensin-converting enzyme 2 (ACE2) and uses ACE2 as an entry receptor to invade the host cells [40]. The serological immunoassays (IA) evaluate the presence of IgM and IgG, produced by the adaptive immune response to SARS-CoV-2 exposure. IgM arises first following a SARS-CoV-2 infection, peaking at week 2-3 while high-affinity IgG rises on week 5-6 and persists for a more significant time. A serosurvey provides epidemiological insights on the prevalence of exposure to the virus. To increase tracking of infection rates, improve epidemiological models, and extend testing capacities beyond central laboratories, CPT code 86328 was approved for a single step “Immunoassay for infectious agent antibody(ies), qualitative or semi-quantitative, single-step method (*e*.*g*., reagent strip).” As of May 11, 2020, the FDA has granted emergency use authorization (EUA) to six point-of-service tests including the qSARS-CoV-2 IgG/IgM Rapid test from Cellex, Vitros Immunodiagnostic Products Anti-SARS-CoV-2 Total Reagent Pack (with Calibrators) from Ortho Clinical Diagnostics, DPP COVID-19 IgM/IgG System from Chembio Diagnostic Systems (requires DPP Micro Reader), Anti-SARS-CoV-2 Rapid Test from Autobio Diagnostics, and the COVID-19 ELISA IgG Antibody Test from Mount Sinai. Abbott Laboratories also announced plans to launch and scale a lateral flow IgG antibody test.

The results of this investigation may have immediate clinical, economic, and societal relevance as an aid in developing antibody testing and return-to-work criteria. The strengths of this report include conversations with clinical experts and survey data from >85 clinicians and epidemiologists. Limitations of our evaluation of antibody testing include sparse validation data of the technology, concern over potential false positives due to cross-reactivity with other human coronaviruses (HCoV-229E, NL63 or OC43), SARS, or MERS, and a lack of published data on the relationship between SARS-CoV-2 antibody positivity and protection. Negative results may not rule out SARS-CoV-2 infection. Simulation exercises on sensitivity and specificity metrics needed for herd immunity in the population were proposed, but modeling results was not available as of April 6, 2020. Future directions by the Diagnostics Accelerator include a detailed evaluation of DTC tests. Large-scale, longitudinal population-based validation studies with data on symptom-free days and contact tracing are needed better to characterize the accuracy of DTC antibody tests for SARS-CoV-2, to understand the prevalence of SARS-CoV-2 exposure among healthcare workers and the general population and elucidate the duration of protection.

Our aims for the horizon scan were two primary objectives: (1) to establish a comprehensive list of currently available COVID-19 tests with the potential to be administered in a DTC setting; and (2) to develop a framework for filtering and ranking tests, to identify top candidates for further study (both now and in the future, as tests emerge). First, we assembled a team of investigators to identify potential test candidates, develop a scoring methodology by which to assess them, and complete both secondary research and direct deep-dive outreach for information. Information on the availability of high-performance COVID-19 tests is rapidly evolving. To ensure the continued use of this evaluation framework for potential DTC test candidates, we have formatted our search results and scoring algorithms in a live, embeddable spreadsheet, publicly available at Mass General Brigham Center for COVID Innovation website (https://covidinnovation.partners.org/diagnostics-direct-to-consumer/). We hope that this framework will prove useful for other institutions looking to establish a method for evaluating potential DTC COVID-19 tests.

The goal of this work was to identify high-performing, rapid testing solutions to deploy at the necessary scale to allow the economy to be safely re-opened. In addition to performance metrics, manufacturing capacity, and supply chain assessments, it will be essential also to consider the ease of procurement when considering which tests to use in the community as there was a significant difference in acquisition time (ranging from a couple of days to several weeks) between products. Commonly, lengthy acquisition times were due to foreign products being delayed by US Customs. However, some instances were related to slow response times by the companies. These factors will need to be accounted for when selecting products for deployment.

### Engagement Protocol

The completion of the evaluation phase of the process resulted in a winnowed group of technologies and providers of interest to explore in more detail. It is helpful to consider several aspects of this more detailed exploration before engaging those providers selected in the evaluation phase. Questions not necessarily covered during the evaluation process and of interest before an engagement includes:

Does a confidentiality agreement (CDA) exist between the evaluator and the provider? Will a CDA be necessary before further information being transferred? The evaluator should consider whether there will be confidential information exchanged in both directions or just from the producer to the evaluator. This will form the basis of a one-way versus a two-way CDA. The evaluator should carefully consider the creation of a CDA and the impact it may have on future data sharing and publication of the evaluation. Should an agreement exist, disclosure of data to third parties not covered by the CDA will not be possible. Assuming materials will be transferred to the evaluator during the next phase, what form will an agreement take to facilitate that transfer? The evaluator commonly takes two paths:

The purchase of materials from the provider in small quantities is sufficient for further analysis. It is common in this case that, following the purchase, the evaluator will have no additional obligations to the provider. The evaluator should carefully review the purchase and sale agreement to avoid language regarding the use of the technology leading to a restriction of the evaluation or publication of results. The receipt of materials from the provider at no cost subject to agreed-upon obligations on the part of the evaluator regarding the management of information obtained during this more detailed examination. Such obligations can be included in an Institutional Service Agreement, Materials Transfer Agreement, Collaboration Agreement, Sponsored Research Agreement, or other mechanism suited to the purpose. Discussions with the Office of General Counsel will be a valuable part of establishing the agreement type most appropriate to the details of the evaluation.

It is rarely the case that new intellectual property will be created during an evaluation of an existing product. The evaluator should consider this potential and either avoid performing work that could lead to new inventions or negotiate intellectual property terms with the provider. Such terms would include, but not be limited to, assignment of rights, option to license rights, future royalties on the use of the intellectual property.

### mHealth Solutions

The mHealth technologies that we examined as part of the study have great potential for providing valuable information for the identification of COVID-19 cases. ePRO platforms have a clear role in identifying individuals reporting symptoms that are consistent with COVID-19. Testing would follow the report of symptoms to confirm or rule out a COVID-19 diagnosis. However, a portion of the COVID-19 population does not experience severe symptoms. There have been even reports of COVID-19 patients who were asymptomatic. Importantly, patients are pre-symptomatic for a relatively long time. The use of ePRO platforms to collect self-reports of symptoms is meaningless in these cases. Preliminary evidence suggests that it is possible that at least a portion of the COVID-19 population would display changes in physiological variables before they report symptoms. Wearable sensors examined in this study would have the capability of detecting deviations from normative values of such parameters. As knowledge about the disease is growing, and new symptoms continue to emerge from clinical reports, it is likely that the crucial physiological to achieve early detections of infections will emerge. Still, evidence that wearable sensors could augment the information gathered using ePRO platforms is limited. Finally, electronic-based contact tracing could provide an essential complement to the information collected using ePRO platforms and wearable sensors. Unfortunately, this technology has its limitations. For instance, contact tracing smartphone apps are typically based on exchanging encrypted identifiers with smartphones within the Bluetooth radio range. Unfortunately, this technique cannot easily distinguish proximity from physical contact, which is what leads to a viral transmission. Besides, smartphone apps are typically not designed to detect viral transmission via fomites (*e*.*g*., apps are unable to determine if an individual used the same seat in a subway car that a COVID patient used a few minutes earlier).

Nonetheless, the information gathered by the mHealth Working Group that we assembled as part of the DTC group of the MGBCCI supports the statement that mHealth technology, as a while, could have a significant role to play in preventing and/or mitigating the effects of the COVID-19 pandemic. Specifically, the technology has the potential to provide relevant information to the assessment of the health risk both at the individual and at the societal level. In individuals who have not been tested, information gathered using mHealth technology could be utilized to determine the probability of infection and hence develop appropriate strategies to deploy diagnostic and immune status tests, including the DTC tests discussed in this manuscript.

A second consideration that emerged from the work of the mHealth Working Group is the relevance of further integrating mHealth technologies into the “regular” clinical practice. This is an ongoing process in many healthcare networks. Because the integration of mHealth technology into clinical practice and protocols has not been fully achieved as yet in many healthcare networks, using such technology in response to COVID-19 is more challenging and risks to disrupt the clinical workflow. Once the integration of mHealth technologies into clinical practice and protocols is accomplished, the process of redirecting these resources to address a pandemic would be simply defined as part of the work that many healthcare networks would carry out to prepare for pandemics and disaster situations.

Finally, mHealth technologies are designed to gather a massive amount of data, but truly what we are interested in is to estimate the health risk of the individual and the community. Deriving such information from data collected using mHealth technologies is not a trivial task. It would likely need to rely on sophisticated data analytics to be validated *via* extensive data collections aimed to provide a solid demonstration of the feasibility of using such information to keep us safe.

### Combining Molecular, Antigen and Serological Tests with Collected Data Using mHealth Technology

Whereas molecular and serological tests, as well as mHealth technologies, have limitations and could not be used effectively to address the COVID-19 pandemic if taken alone, an interesting question is whether their use can be combined in a way that maximizes the number of detected cases while deploying reasonable resources. Figure 6 schematically represents the approach that we propose. In this conceptual schema, individuals in the community would first be asked to use a smartphone app or equivalent to report symptoms consistent with COVID-19. If symptomatic, they would immediately undergo diagnostic testing. If not symptomatic, they would undergo immune status testing. Besides, a random sample of individuals will also be instructed to undergo diagnostic testing in an attempt to capture those who are infected but present with no symptoms. Among those who are positive to the immune status test, a subset would be directed to explore if they can serve as donors for plasma therapy, a portion would be randomly selected for diagnostic testing and monitoring to gather data concerning immune protection, and the remaining individuals would undergo home monitoring using mHealth technology. Instead, those who are negative to the immune status test would all undergo home monitoring using mHealth technology. Monitoring would consist of periodic self-reports of symptoms, physiological data collected using wearable technology, and the use of smartphone apps for electronic contact tracing. Clinical data related to risk factors relevant to COVID-19 would be gathered. Also, data concerning the health status of family members and others in close contact with the monitored individuals would be tracked if possible. All these data would be fed to a probabilistic model that would continuously assess the probability of infection. Individuals whose data suggests a high to moderate probability of infection would be directed to undergo diagnostic testing either at home or a at a central lab. Standard medical procedures would be adopted for those who test positive for COVID-19. Individuals whose data suggests a very low probability of infection, as well as those who test negative for COVID-19, will go back into the pool of subjects undergoing home monitoring and a subset would be randomized to undergo immune status testing, again, in an attempt to capture subjects who might have been infected but did not experience any symptoms.

**Figure 6.**
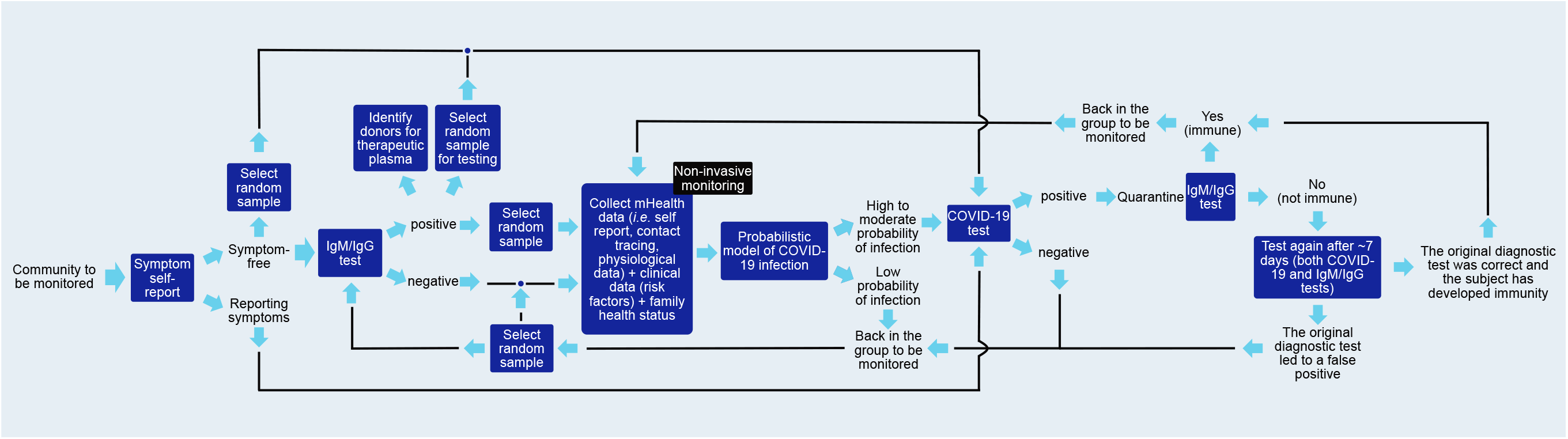
Conceptual schematic framework represents combining molecular and serological tests with data collected using mHealth technology.

Whereas the schematic shown in Figure 6 provides a general description of the proposed approach, many detailed aspects of the implementation of such a plan would need to be addressed. For instance, one would need to determine criteria for sampling the population of those symptoms free to explore ways to detect individuals who have been infected but present with no symptoms. Strategies such as repeating tests and resampling the community undergoing tests would need to take into account data concerning false positives and false negatives associated with each testing methodology. Importantly, the choice of mHealth technologies to be used to collect data in the home and community settings would need to be revisited as our knowledge of the disease and associated physiological response improves. Finally, the approach to build a model to process the data collected in the home and community would need to be chosen. Data collected from tests would need to be used as labels to train machine learning algorithms part of such models to generate accurate estimates of the probability of infection on an individual basis.

## Conclusions

Recent advances in molecular and serological testing provide health care officials with the opportunity to implement a massive testing campaign based on direct-to-consumer products to monitor and prevent COVID-19 infections. Available testing techniques for home-based testing are far from perfect. Still, their use could be complemented by data collected using mHealth technology to monitor the health status of individuals in the community. In this manuscript, we propose an approach that would combine these two tools (molecular and serological testing with data collected *via* monitoring with mHealth technology) to maximize the likelihood of rapidly detecting COVID-19 cases and individuals who have been in contact with COVID-19 patients to minimize the consequences of the pandemic.

## Data Availability

The datasets generated during and/or analyzed during the current study are available at the Mass General Brigham Center for COVID Innovation (MGB-CCI) website.

https://covidinnovation.partners.org/diagnostics-direct-to-consumer/

## Acknowledgments

We are indebted to the volunteers for their participation in the MGB CCI Direct-To-Consumer Task Force and working group as well as the investigators and staff at each of the working groups of the MGB COVID Innovation Center who made this investigation possible.

## Author Contributions

A.H., M.J., and F.H. led the Specifications, N.M.L. led the Horizon Scan, F.H. led the IRB, P.B. led the mHealth Working Group, G.M. led the Engagement and Solution Rollout, M.J. led the data and publication, J.A.T. led the program office, R.A. led the DTC working group, and D.R.W. co-led the MGB Center for COVID Innovation and led the Diagnostics pillar. M.J. wrote the manuscript with contribution from all authors.

## Notes

### Competing Interest Statement

The authors have declared no competing interest.

### Funding Statement

No external funding was received.

### Author Declarations

No IRB is needed for this study

